# Crossing the digital divide: The workload of manual data entry for integration between mobile health applications and eHealth infrastructure

**DOI:** 10.1101/2024.04.23.24306024

**Authors:** Caryl Feldacker, Joel Usiri, Christine Kiruthu-Kamamia, Geetha Waehrer, Hiwot Weldemariam, Jacqueline Huwa, Jessie Hau, Agness Thawani, Mirriam Chapanda, Hannock Tweya

## Abstract

**Background:** Many digital health interventions (DHIs), including mobile health (mHealth) apps, aim to improve both client outcomes and efficiency like electronic medical record systems (EMRS). Although interoperability is the gold standard, it is also complex and costly, requiring technical expertise, stakeholder permissions, and sustained funding. *Manual data linkage* processes are commonly used to “integrate” across systems and allow for assessment of DHI impact, a best practice, before further investment. For mHealth, the manual data linkage workload, including related monitoring and evaluation (M&E) activities, remains poorly understood.

**Methodology:** As a baseline study for an open-source app to mirror EMRS and reduce healthcare worker (HCW) workload while improving care in the Nurse-led Community-based Antiretroviral therapy Program (NCAP) in Lilongwe, Malawi, we conducted a time-motion study observing HCWs completing data management activities, including routine M&E and manual data linkage of individual-level app data to EMRS. Data management tasks should reduce or end with successful app implementation and EMRS integration. Data was analysed in Excel.

**Results:** We observed 69:53:00 of HCWs performing routine NCAP service delivery tasks: 39:52:00 (57%) was spent completing M&E data related tasks of which 15:57:00 (23%) was spent on manual data linkage workload, alone.

**Conclusion:** Understanding the workload to ensure quality M&E data, including to complete manual data linkage of mHealth apps to EMRS, provides stakeholders with inputs to drive DHI innovations and integration decision making. Quantifying potential mHealth benefits on more efficient, high-quality M&E data may trigger new innovations to reduce workloads and strengthen evidence to spur continuous improvement.

## Introduction

In sub-Saharan Africa (SSA), mobile health (mHealth) innovations are expanding rapidly. mHealth tools are recognized by the World Health Organization (WHO) for their potential to improve the quality of client care across diverse health sectors and healthcare contexts ^1^. Digital health interventions (DHIs), like mobile health (mHealth) apps, may require less consistent electricity or connectivity; be faster to design; and lower costs over traditional electronic health (eHealth) systems ^2-4^. mHealth also holds promise to extend the reach of traditional electronic medical record systems (EMRS) that typically operate in static hospitals and health facilities. Despite high development and maintenance costs^5^, EMRS are widely recognized to improve client outcomes via provider decision making supports that promote adherence to complex clinical guidelines, including those for antiretroviral therapy (ART). EMRS also manage high data volumes, which can streamline critical monitoring and evaluation (M&E) data for reporting and increase efficiency. To support improved M&E of client and program outcomes in line with global guidance ^6^, Ministry of Health (MoH) supported EMRS are scaling nationally across many low and middle income countries (LMICs) ^7^.

The implementation of both DHIs and EMRS brings benefits to clients, providers, and programs. Yet, many DHIs, including mHealth, largely operate without links to MoH supervision, routine data aggregation pipelines, or national M&E reporting systems ^2,3,8^. Operating outside of national eHealth infrastructure may reduce potential impact, replication and scale while also undermining MoH authority. Data fragmentation between bespoke apps and EMRS also reduces client care quality, breaking the flow of M&E information needed to manage clients effectively and efficiently along the continuum of care ^9^. Lack of integrated data between systems, and appropriate policy to govern and coordinate efforts at the national level, also reduces M&E data quality for MoH decision-making, potentially reducing sufficient resource allocation to attain improvements in population-level outcomes ^10,11^.

Interoperability or integration between digital health tools like mHealth and EMRS is considered the gold standard to ensure complete, correct, and consistent bi-directional data flow between distinct healthcare delivery settings and systems. Across SSA, there is commitment to, and momentum for, alignment between digital health systems and larger, MoH-driven e-health policy and practice ^12^, including efforts to support common client registries. Several operating frameworks provide guidance on the complex process of system unification between EMRS and DHI deployments, including adoption of organizational standards, architecture requirements, legislation or governance recommendations, standardised data elements, and protections of client privacy ^13,14^.

Although definitions of interoperability and integration vary widely and are not well standardized ^15-20^, both concepts include bi-directional data exchange. *Integration* may be considered a lower technical bar that employs a customized middle layer to translate data between systems. Integration may be more appropriate for highly customized, local, or national EMRS. Along the complexity continuum, *interoperability* typically requires a higher technical standard, often characterized by centralized data repositories and a unified system of digital data management like open health information exchange (OpenHIE) ^21^. Interoperability may require adherence to a common OpenHIE middle layer and globally-recognized standards, such as Health Level 7 (HL7) Fast Health Interoperability Resources (FHIR) ^22^, to facilitate bi-directional communication with broader eHealth infrastructure. Interoperability may be more achievable in mature eHealth contexts where EMRS already comply with international standards ^15^. For the purposes of this paper and for simplicity, we use the singular term, *integration*, to reflect an automated, bi-directional communication channel between mHealth and EMRS.

Despite established benefits and global support, integration between mHealth innovations and existing EMRS operating at scale faces formidable challenges. Integration is complex and costly, requiring multi-level buy-in, technical expertise, adequate training and mentoring, stakeholder permissions, and sustained funding ^23^. To be effective, integration requires local adoption, optimization, and multi-level approval ^24-27^, processes that are both critical and cumbersome. Integration between mHealth and EMRS may not be recommended nor feasible in all cases or contexts. For example, some mHealth apps may fail to achieve their objectives ^28-30^; have distinct, short-term utility (i.e., COVID testing); are employed only in research or pilot settings ^31^; or target a highly specific group of users (i.e., orphans or vulnerable children) – all cases where integration may not be advisable nor achievable ^28,30,32,33^.

Before integration investment, and in alignment with digital health best practices^34,35^, mHealth apps should first provide clear evidence of success according to agreed upon, *a priori*, individual-or program-level benchmarks. Apps should demonstrate consistent, correct, complete, and high-quality data for M&E, providing assurances to MoH stakeholders that app data aligns with local and national client and program M&E data standards, before establishing integration with existing, successful EMRS. During this phase, *manual data linkage*, where healthcare workers (HCWs) enter client M&E data by hand to “integrate” across systems and facilitate the continuum of client data between services or service delivery points, may be advisable while rigorous mHealth impact evidence is gathered and data quality considered by stakeholders. When completed well, this manual linkage of client-or aggregate-level data between systems may address several common integration challenges, including delays in eHealth decision-making across many players/politics; frequent data pipeline updates; or, lack of reliable connectivity to transfer data. Manual data linkage workload and its costs are likely common to bridge mHealth and EMRS integration, yet they remain poorly understood.

Therefore, to better understand the costs of this intermediary step before integrating mHealth with an EMRS, we undertook a time-motion study of the manual M&E processes required to link data from an mHealth app with the MoH ART-focused EMRS in Lilongwe, Malawi. We included M&E workload as reducing workload is a primary objective of the app. The mHealth innovation, the **C**ommunity-based **A**RT **RE**tention and **S**uppression (*CARES*) App, intends to provide EMRS-like advantages for ART clients and for M&E data quality in Lighthouse Trust’s (LT) Nurse-led, Community-based ART Program (NCAP). CARES was designed by the University of Washington’s International Training and Education Center for Health, Lighthouse Trust, and Medic using the open-source Community Health Toolkit (CHT) ^36^. CARES’ functionality and design features reflect the priorities of both the Government of Malawi and US President’s Emergency Plan for AIDS Relief (PEPFAR) ^37^. At the end of the ongoing optimization and evaluation phase, CARES’ aims to deliver real-time advantages for NCAP client clinical care; workload reduction for providers and M&E-focused HCWs; and benefits in M&E data quality and access resulting from integration with the static site EMRS.

Our objective in this baseline time-motion assessment is to describe the workload of data-related tasks, including manual data integration between the open-source CARES app and the customized Malawi EMRS, to understand the potential workload reduction from successful app launch and EMRS integration. This embedded costing activity is part of an ongoing implementation science study of CARES app effectiveness. As both routine M&E and manual data exchange processes are likely not unique to CARES nor for other mHealth deployments, we hope these findings and may inform future decision making for others using open-source apps with aims to integrate into existing EMRS infrastructure.

## METHODS

### Study design

We conducted a descriptive, time-motion costing study observing the workload of 7 NCAP nurses, 2 M&E data officers, and 4 other providers (1 retention assistant, 1 ART clerk, 1 Expert Client (an ART support services healthcare cadre), and 1 community care supporter) as they completed NCAP related activities and data management tasks (Table 2). The perspective is from the HCW workload. As with many routine HCWs, some HCWs performed other routine duties outside of NCAP related tasks, but were observed during discrete periods of NCAP tasks. Therefore, observations were collected and considered proportionally: total observed hours of NCAP-related activity across HCWs were categorized to estimate the proportion of NCAP-related time spent on service delivery unrelated to data management, CARES-related tasks or EMRS <> CARES-integration tasks. This pre-CARES time-motion study serves as a baseline for future repetition of the time-motion study, post CARES implementation.

### Setting

LT is an MoH service provider in Lilongwe, Malawi that provides integrated HIV care to 38,000 ART clients in its two flagship clinics in urban Lilongwe: 26,000 at Martin Preuss Centre (MPC) and 12,000 at Lighthouse clinic (LH). Like other large Malawi ART clinics, LT employs a real-time, point-of-care (POC) EMRS to ensure adherence to MoH integrated ART guidelines and ease routine M&E reporting EMRS^38^. The highly customized EMRS scaled nationally across Malawi ^39,40^, supporting quality client care, provider decision-making, and national reporting indicators in over 600 large, MoH ART facilities ^41^.

### Description of NCAP services

LT operates a large-scale, client-centered, differentiated service delivery (DSD) model: the nurse-led community-based ART program (NCAP) ^42^. In NCAP, nurses provide ART services to stable LT clients every 3-6 months through peer support meetings. The NCAP serves 2400 clients from two central hospitals and 2800 clients from rural/peri-urban satellite sites across 120 groups in Lilongwe District. NCAP nurses deliver ART, conduct rapid clinical reviews, draw viral load (VL) blood samples and deliver lab results in the community. NCAP is widely supported by providers and clients, alike. Unlike in static sites, NCAP nurses do not have access to the static LT EMRS, potentially reducing the quality of integrated client care and increasing the M&E workload. NCAP client visits are documented in tablets using Open Data Kit (ODK) data management forms. ODK spreadsheets are printed for manual EMRS entry by clerks. ODK has several limitations including lack of clinical decision support to HCWs ^43^. Clerks enter data each week; data is verified by routine M&E checks before appending to EMRS datasets for inclusion in routine MoH reporting.

### Description of the CARES app

The Community-based ART REtention and Suppression (CARES) app was designed in 2022 via an iterative, highly-participatory, human-centered design (HCD) process (Table 1) ^43^. CARES local specification and optimization reflects inputs from client, HCW and MoH stakeholders. To reduce design costs and increase potential scalability, CARES leveraged an open-source, global good: the CHT. CARES was built on the existing CHT core framework. CARES implements core characteristics of mHealth best practice, including optimizing HCW accessibility and acceptance; aiming to lower costs, prioritizing local adaptation, building strong stakeholder collaboration, and establishing government partnership for sustained impact ^44^. During NCAP visits, CARES leads nurses through a complete ART review, mirroring the modules, question flow, embedded prompts and decision supports from EMRS. To date, integration between CARES and EMRS has not been established. Complete visit data from the CARES app is printed for manual entry into the EMRS, parallel to the NCAP data management process, as the CARES app is launched, tested, improved, and optimized. Anticipated M&E gains and workload reduction from CARES is presented in Table 2.

**Table 1:** Anticipated benefits of CARES mHealth app on community-based care.

| Activity | Current NCAP | Anticipated CARES Value-add for NCAP |
| --- | --- | --- |
| Client verification during follow-up | Nurse enters client ID into tablet to search a client. | Nurse scans client barcode, confirms client ID, and pulls previous visit data for follow-up |
| Patient treatment history | No previous treatment data available | Previous information on latest VL,TB, side effects, and family planning methods |
| Complete client assessment | Abridged assessment, adherence | Integrated services and care alerts on clinic review (e.g., annual VL due, hypertension, Family planning, TB) |
| Daily NCAP drug management | Nurses pull paper files for expected clients. Collect and manually reconcile ARV drugs at ARV pharmacies | Digitized collection forms to improve M&E efficiency and reduce data errors |
| NCAP ART to EMRS data link | None. Data from tablet is manually entered into EMRS | Offline CARES data synced to site EMRS using client IDs. M&E workload streamlined, reducing costs. |

**Table 2:** CARES expected impact on streamlined M&E workload and costs.

| Table 2: CARES expected impact on streamlined M&E workload and costs |  |
| --- | --- |
| Activity # | Definition of activity |
| Category 1: Nurse tasks not expected to benefit from CARES |  |
| 1 | Registering client in NCAP program |
| 4 | Request and collect ARVs, CPT, INH and Pyrodoxine for clients appointed at the outreach clinic from CHS office |
| 6 | Provide integrated ART- provide ARVs, vital sign, hypertension, TB, family planning services, collect and management viral load samples, Document start time of ART services and end time. Document number of times of clients seen. |
| 12 | Other including transporting medical supplies and equipment, drug collection from CHS office, etc. |
| Category 2: Potential benefit from CARES App |  |
| 2 | Generate list for NCAP clients who are due for ARV pick up in the distribution points |
| 3 | Reconciliation of previous supplies to be eligible for the new supplies. Request and collect ARVs, CPT, INH and Pyrodoxine for clients appointed at the outreach clinic from respective health facility pharmacy |
| 5 | Reconcile all drugs and supplies collected after NCAP visit |
| 9 | CHS nurse generates of list of clients who missed the NCAP visit. Currently, this potential lost to follow-up (LTFU) list is created manually |
| 10 | Prepare the list of clients who submitted VL samples for each site, Update VL results received in the month, |
| 11 | Prepare daily, weekly, monthly and quarterly NCAP reports |
| Category 3: Potential benefit from CARES Integration |  |
| 7 | Download, filtering, and printing from ODK files |
| 8 | Enter data retrospectively with vital signs, drug dispensed, next appointment date from Lighthouse NCAP database into the facility EMR, Print visit summary and stick them on clients charts in collaboration with receptionists |

### Description of routine monitoring and evaluation (M&E)

Routine M&E includes tasks related to the process and transformation of data for decision making at the client, clinician, clinic, and aggregate levels. In the NCAP and CARES context at LT, this includes: tasks related to gathering EMRs data for expected NCAP clients, including those requiring VL samples; creation of a paper list of clients who missed expected NCAP visits; data downloading and printing from the app; performing routine verifications of M&E data quality before EMRS uploading; and manual data entry of printed into the EMRs HCW compile routine NCAP reports. Once entered in the EMRS, NCAP data from ODK or from CARES is included for all routine MoH M&E reporting, requiring no further or future modification.

### Definitions

Routine HCWs from NCAP and M&E teams determined the list of HCW activities that would be included in the observation check-list and categorized tasks into three groups (Table 2): 1) *Not Related* are routine NCAP activities that are not expected to benefit from CARES app implementation; 2) *Benefit from CARES App* are NCAP activities that are expected become more efficient with CARES implementation, independent of linkage to the EMRS; and 3) *Benefit from CARES Integration* are NCAP activities that are expected to require little or no time once CARES is integrated into the EMRS. Observed tasks included those that were both directly and indirectly associated with NCAP data collection using CARES.

### Data collection

To facilitate time tracking, LT research assistants (RAs) performed activity timekeeping, observing different cadres of LT routine HCWs along each step of the NCAP data pathway, including NCAP file preparation, community-based data collection, data management, and manual entry into the EMRS. Not all activities happen on all days. The RA used the stopwatch function on their phone to record the time required for staff to complete their tasks. For each activity, the number of clients was estimated and facilities recorded. Observations of the 13 providers were made over 17 days of NCAP service delivery, including tasks related to CARES data collection and other activities (6 observation days in Oct 22; 11 observation days in May-June 2023). Observations included both routine and periodic activities using the same checklist and timekeeping methods. Manual data linkage occurred on 7 of the 17 calendar days where observations occurred. As with the organization of routine NCAP and LT operations, some HCWs work, and were observed, performing multiple tasks in multiple facilities on the same day.

### Data analysis

Of the 120 activity observations, 48 observations were related to activities 1, 4, 6, and 12 --activities that were not expected to be affected by the CARES app or CARES integration. The remaining 72 observations were of activities 2, 3, 5, 7, 8, 9, 10, and 11, highlighted in Table 1.

#### Total time

Time spent on each observation was calculated as the difference between activity start and end time. Time spent on each CARES-related activity was calculated by summing times across observations for each activity. We calculated time by location, by provider type, and total time across all selected activities. We divided total CARES-related time into two CARES-related categories – time on activities affected by the CARES app and time spent on CARES integration activities. Data was analysed in Excel.

### Data Availability statement

Data will be made available at Dryad upon acceptance at this link: https://doi.org/10.5061/dryad.k0p2ngfdz Participant initials and exact dates for data collection were replaced with pseudonyms and approximations in the shared dataset.

#### Ethics

The CARES study protocol, including this costing component, was approved by the Malawi National Health Sciences Research Committee (Protocol# 21/11/2830) and the University of Washington, Seattle, USA (STUDY00013936) ethics review board. HCWs verbally consented for observations completing routine work in routine settings. No identifiable data was collected for this costing study.

## RESULTS

### Participant and observation characteristics

Thirteen HCWs across five facilities were observed during the time-motion study (Table 3). Of the 120 activity observations, 72 activities were expected to benefit from CARES implementation, including integration (Table 3). The teams were all highly qualified. All nurses were Community Health Nurses. HCWs at Lighthouse, Kawale, and Mitundu typically had an academic qualification of a diploma or above, while at Chileka and Nathenje, HCWs more commonly had a Junior Certificate of Education. Most had 1-5 years experience at their position, but several had more than ten years of experience at their jobs.

**Table 3:** Participant and observation characteristics, by facility.

| Facility Name | Cadres Observed* | # observed activities | # CARES-related observed activities |
| --- | --- | --- | --- |
| Lighthouse | Data officer, nurse | 94 | 57 |
| Chileka | Other | 1 | 1 |
| Mitundu | Nurse, other | 16 | 7 |
| Kawale | Other | 1 | 1 |
| Nathenje | Other, nurse | 8 | 6 |
| Total |  | 120 | 72 |

\*other includes: ART clerks, expert clients, and retention assistants

### Workload by workload type

Total observation time related to NCAP services was 69 hours and 53 minutes (Table 4). HCWs spend an average of 39 hours and 52 minutes (57% of total time) to complete data-related tasks, of which 23:55:00 was related to tasks targeted by CARES efficiency gains and 15:57 related to manual data integration tasks. Among all observed workload, providing integrated ART took the most time (27 hours and 59 minutes), a non CARES-related task. The second largest amount of time was spent on a CARES-related M&E activity, generating a list of potential lost to follow up clients, with over 9 hours or 13% of all observed time (40%). Tasks related to integration took 23% of all observed HCW time spent on NCAP.

**Table 4:** Time spent by NCAP activity, by activity type.

| **Activity #** | **Count of activities observed** | **Time spent on activity** | **Percentage of overall time** |
| --- | --- | --- | --- |
| Category 1: Nurse tasks not expected to benefit from CARES |  |  |  |
| 1 | 0 | 0:00:00 | 0% |
| 4 | 4 | 0:39:00 | 1% |
| 6 | 23 | 27:59:00 | 40% |
| 12 | 21 | 1:23:00 | 2% |
| Category 2: Benefit from CARES App |  |  |  |
| 2 | 3 | 0:38:00 | 1% |
| 3 | 1 | 1:20:00 | 2% |
| 5 | 1 | 2:37:00 | 4% |
| 9 | 20 | 9:09:00 | 13% |
| 10 | 19 | 7:01:00 | 10% |
| 11 | 12 | 3:10:00 | 4% |
| Category 3: Benefit from CARES Integration |  |  |  |
| 7 | 7 | 8:34:00 | 12% |
| 8 | 9 | 7:23:00 | 11% |
| **Total** | **120** | **69:53:00** | **100%** |

### Workload by Cadre

When stratified by provider cadre, Community Health services nurses took up the most amount of time on CARES related activities-25 hours and 14 minutes (Table 5), followed by data officers at 10 hours and 19 minutes. Data officers and other HCWs (ART clerks, expert clients, and retention assistant) spent all or most of their time on CARES integration activities while community health nurses spent the least time on these activities.

**Table 5.**
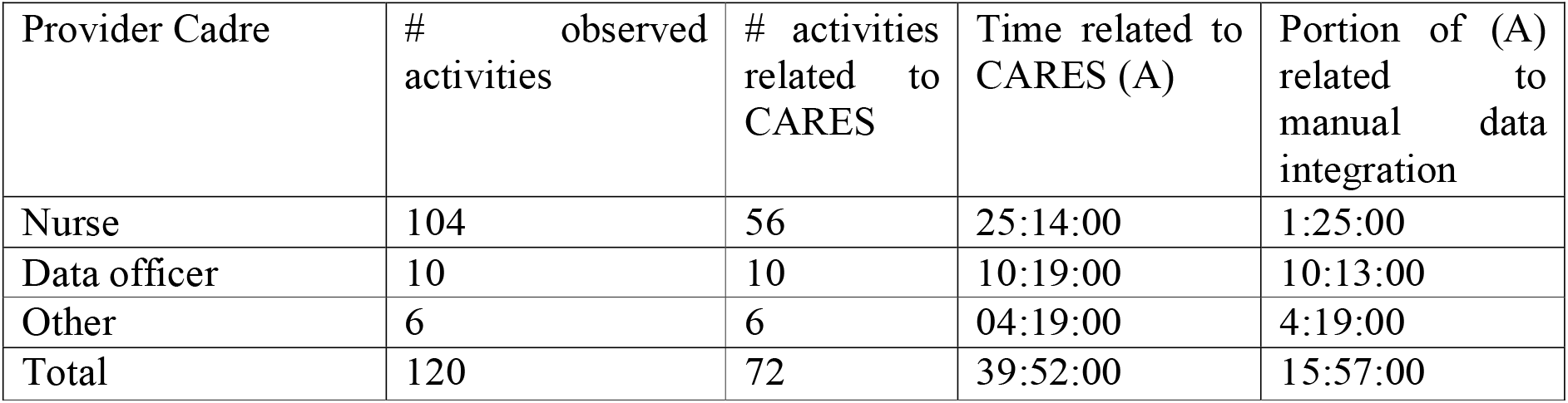
Time taken for CARES-related activities stratified by participant cadre.

## DISCUSSION

In this focused time-motion study, we observed almost 70 hours (69:53:00) of NCAP-related service delivery and found that HCWs spend an average of 39 hours and 52 minutes (57% of total time) to complete M&E data related tasks. Of the 39:52:00 collecting or managing data, 23 hrs 55 mins, or ~60% of all data-related time, could be reduced by CARES app use, itself. Although the goal of CARES and many other mHealth apps is to contribute to a common digital infrastructure that is interoperable with the EMRS, this was not possible in the short term. In the current Malawi e-health environment ^45^ and CARES prototype stage, manual data exchange between the mHealth app and the EMRS is a digital health best practice to allow for app optimization and rigorous testing while ensuring the continuum of timely, complete, M&E data to optimize client and program outcomes. The required resources to complete this critical step along the pathway to integration are not negligible. The manual data linkage process to ensure the continuum of client data between the mHealth app and the Malawi EMRS, took approximately 15 hours and 57 minutes, 40% of the data-related workload or ~23% of all HCW time providing NCAP services. If successful, it is expected that CARES app and integration with EMRS will considerably improve efficiency, reducing M&E workload. By quantifying the impact of mHealth programs on M&E workload, we can strengthen justification to continuously invest in mHealth improvements. Our findings have several implications for other mHealth apps using manual data linkages along the pathway to integration.

First, for CARES and for mHealth implementation, allocating sufficient resources to manual data exchange between mHealth and EMRS while an app is rigorously evaluated allows time to address poor app performance, data quality concerns, or operational inefficiencies that could inhibit successful integration across systems. Pushing DHI integration with EMRS before data quality assurances and positive app impact are established may significantly threaten the quality of EMRS data; compromise patient personal identifying data; or be wasteful in the context of innovation research where digital health interventions may fail. Although some national EMRS systems use open-source platforms which can facilitate mHealth tool integration ^26,46,47^, the integration process for many DHIs is still lengthy and resource-intensive^9^. Similarly, use of global, open source, mHealth tools like the CHT on which CARES was built, may also facilitate interoperability with standardized e-health tools, but face greater hurdles with highly customized EMRS, like in Malawi, that were not designed for interoperability. Interoperability in all contexts requires complex engineering that may be out of reach for many LMIC mHealth implementers with typical tight timelines and budgets. Moreover, with constantly adapting eHealth systems, integration as a singular threshold or achievement may also be inaccurate. As eHealth systems change and apps are optimized, manual data exchange processes may be intermittently required as a stop-gap. Planning for this eventuality could ensure sufficient resources to ensure quality M&E data throughout the mHealth to EMRS integration pathway.

Second, we demonstrate that the M&E workload, including tasks associated with manual data linkage, is a significant burden for HCWs. Quality M&E data is critical for routine and periodic client and program assessment. Routine M&E, especially tasks associated with data linkage between systems, must be undertaken by well-trained HCWs and supported by quality assurance measures to ensure data quality. Costing studies, including those in mHealth, often neglect the M&E workload, missing critical components of routine work such as data preparation, collection, transformation, entry, and quality verification processes. In this study, we intended to uncover the costs of M&E tasks along the app-to-EMRS integration pathway that may be missed in other costing studies, including the time and resources needed for retrospective data entry workload; activity planning and management; stock reconciliation; and routine ART program M&E reporting in line with MoH requirements. Quality M&E data is not only needed to assess mHealth impact on various facets of service delivery, but is required by many MoH, including in Malawi ^48^, as well as the global donor community ^49^. Our findings also suggest that mHealth should consider targeting potential M&E efficiency gains in routine service delivery settings-with benefits that could foster cost reductions beyond those typically considered for client, frontline HCWs, and program perspectives. Savings in client-and program-level M&E tasks, as a result of quality DHI, could translate to increased available resources for investment in integration.

Several aspects of the workload costs incurred at Lighthouse and within the Malawi integration context may be different than those in other routine LMIC settings, including in SSA, and should be considered for future costing. For example, in any setting clerks must be highly trained and supervised to ensure quality data is entered into the EMRs to avoid inconsistent or incorrect data for decision making ^45^. At Lighthouse, specifically, clerks had more than 5 years of experience, potentially making these HCWs distinct from many other LMIC contexts where these tasks could take far more time. For M&E tasks, literacy and digital literary skills are required ^46^: this is a common challenge in settings with already scarce HCW resources. At Lighthouse, this issue is not as acute as clerks have at least a high school diploma. Moreover, not all clinics in all settings comply fully with MoH M&E data quality and reporting guidelines. At Lighthouse, M&E data officers undertake routine data quality audits to identify and rectify data errors, requiring additional resources for data collected from either manual or electronic data sources. Not all settings would match the clinic operations at Lighthouse, which may reduce the potential generalizability of these findings. However, in any setting, these additional considerations are important aspects of ensuring high quality M&E data, including for manual data linkage, and merit future consideration in this or other settings.

Lastly, although we highlight the importance of manual data linkage while apps are proven effective and data quality assured, interoperability with national EMRS is still the goal and gold standard for successful DHIs, including mHealth. The adoption of international data standards, like H7FHIR, as completed recently by the Malawi MoH, significantly reduce data sharing challenges and costs within the healthcare systems. Adoption of these standards also gives MoHs tools, guidelines, and requirements to reduce threats from the profusion of DHIs. Interoperability led by MoHs, paired with required governance and compliance regulations, reduces data siloes and creates a holistic view of client information throughout the care continuum. For mHealth designers and implementers, the mHealth community must adhere more carefully, conscientiously, and intentionally to the digital infrastructure requirements of each MoH, including adherence to common client registries or OpenHIE systems required by MoH policy.

## LIMITATIONS

First, unlike traditional time-motion studies, NCAP HCWs do not complete only NCAP related tasks each day; therefore, hours of HCW observation were collapsed over time to give an estimate of the proportion of NCAP-related time, overall, that was used for CARES or CARES-integration. Second, not all facilities contributed data, potentially influenced the average amount of time it took for manual data integration per location. Likewise, not all clerks were observed, suggesting that the average may not reflect all M&E team members in these locations. Given our study was only conducted over 17 seventeen calendar days, our findings may not be sufficient to observe all M&E tasks related to manual data entry nor representative of all potential cost savings from integration. Likewise, we didn’t put a dollar value on workload and salary; the time considerations could be applied to cadres levels in other contexts, hoping to improve generalizability of this small study. Observations were repeated to augment the activity and per-cadre time calculations; changes in the interim in personnel or personnel skills could have increased or decreased time/client calculations. To the extent that these tasks were undertaken by highly skilled nurses and clerks, these estimates may understate the true workload estimate. Lastly, we did not cost out the total costs of CARES app, a costing exercise that is planned for future.

## CONCLUSION

There is clear momentum and support for interoperability between mHealth tools and EMRS. There is also global emphasis on the critical need for high quality, routine M&E data for decision-making to ensure the client care continuum and measure program level impact. Effective mHealth solutions, including those like CARES that aim to extend the reach of EMRS, go beyond improving client care to also generating quality data for program M&E. However, not all mHealth interventions could or should integrate with EMRS at all times. The costs, complexity, and technical skills required for successful mHealth and EMRS interoperability, especially with bespoke EMRS like in Malawi, are high. A first step for mHealth and EMRS integration is appropriate resource allocation for manual data exchange processes to bridge the data divide between apps and existing EMRS, allowing for rigorous DHI evaluation before integration efforts. Assessment of the manual data linkage workload, including routine M&E data tasks, is often neglected in costing studies. Understanding the time and HCW burden associated with M&E tasks that could benefit from mHealth implementation and manual data linkage provides practitioners and policy makers with evidence to drive mHealth decision making. If DHI evaluations can offer evidence of app success, and strongly suggest potential workload reductions in data-related activities, saved costs from mHealth implementation could be invested continuously in integration establishment and maintenance.

## Supporting information

Costing dataset

## Data Availability

All data produced will be available online at Dryad upon publication. Participant initials and exact dates for data collection were replaced with pseudonyms and approximations in the shared dataset.

## Acknowledgments

Research reported in this publication was supported by the National Institutes of Health (NIH), National Institute of Mental Health (NIMH) under award number R21MH127992 (“The Community-based ART REtention and Suppression (CARES) App: an innovation to improve patient monitoring and evaluation data in community-based antiretroviral therapy programs in Lilongwe, Malawi”), multiple principal investigators, Feldacker and Tweya. The content is solely the responsibility of the authors and does not necessarily represent the official views of the NIH nor NIMH. The authors would also like to thank the Lighthouse Trust and its NCAP program for their partnership in the CARES design process.

